# Severity of SARS-CoV-2 Infection in Pregnancy in Ontario: A Matched Cohort Analysis

**DOI:** 10.1101/2022.03.04.22271915

**Authors:** Kiera R. Murison, Alicia A. Grima, Alison E. Simmons, Ashleigh R. Tuite, David N. Fisman

**Affiliations:** Dalla Lana School of Public Health, University of Toronto, Toronto, Ontario, Canada; Centre for Immunization Readiness, Public Health Agency of Canada, Ottawa, Ontario, Canada

## Abstract

**Background:** Pregnancy represents a physiological state associated with increased vulnerability to severe outcomes from infectious diseases, both for the pregnant person and developing infant. The SARS-CoV-2 pandemic may have important health consequences for pregnant individuals, who may also be more reluctant than non-pregnant people to accept vaccination. We sought to estimate the degree to which increased severity of SARS-CoV-2 outcomes can be attributed to pregnancy.

**Methods:** Our study made use of a population-based SARS-CoV-2 case file from Ontario, Canada. Due to both varying propensity to receive vaccination, and changes in dominant circulating viral strains over time, a time-matched cohort study was performed to evaluate the relative risk of severe illness in pregnant women with SARS-CoV-2 compared to other SARS-CoV-2 infected women of childbearing age (10 to 49 years old). Risk of severe SARS-CoV-2 outcomes (hospitalization or intensive care unit (ICU) admission) was evaluated in pregnant women and time-matched non-pregnant controls using multivariable conditional logistic regression.

**Results:** Compared to the rest of the population, non-pregnant women of childbearing age had an elevated risk of infection (standardized morbidity ratio (SMR) 1.28), while risk of infection was reduced among pregnant women (SMR 0.43). After adjustment for age, comorbidity, healthcare worker status, vaccination, and infecting viral variant, pregnant women had a markedly elevated risk of hospitalization (adjusted OR 4.96, 95% CI 3.86 to 6.37) and ICU admission (adjusted OR 6.58, 95% CI 3.29 to 13.18). The relative increase in hospitalization risk associated with pregnancy was greater in women without comorbidities than in those with comorbidities (P for heterogeneity 0.004).

**Interpretation:** A time-matched cohort study suggests that while pregnant women may be at a decreased risk of infection relative to the rest of the population, their risk of severe illness is markedly elevated if infection occurs. Given the safety of SARS-CoV-2 vaccines in pregnancy, risk-benefit calculus strongly favours SARS-CoV-2 vaccination in pregnant women.

## Introduction

Pregnant individuals represent an important priority population for communicable disease prevention, for several reasons. The pregnant state results in changes in the immune system necessary for immune tolerance of the fetus (1, 2), and that may result in greater severity of some infections (1, 3-5). Physiological changes associated with pregnancy, including metabolic and hormonal changes, and mechanical reduction in respiratory reserve, make pregnant individuals more vulnerable to respiratory impairment (5, 6). Management of critical illness in pregnancy is challenging due to the distinct physiology of pregnant people and concerns around the use of some therapeutic agents, while critical illness in the pregnant individual may result in fetal demise (5, 7). Finally, although vaccines in pregnancy, including those that prevent SARS-CoV-2 infection, are safe and effective (8), their uptake may be limited due to aversion to pharmaceuticals on the part of both pregnant individuals and care providers (9, 10). Informed decisions around vaccine acceptance depend on accurate information on the risks of SARS-CoV-2 in the context of pregnancy.

SARS-CoV-2 has infected hundreds of millions of people since the disease emerged in 2019; it has also caused critical illness and death in millions (11). In the United States, pregnant women have been found to be at a markedly elevated risk of critical illness from SARS-CoV-2 infection, as well as an elevated risk of stillbirth (12-14). Initial analyses failed to identify an elevated risk of death among pregnant women with SARS-CoV-2 infection, but a subsequent re-analysis identified increased mortality risk, likely due to increased event numbers with the passage of time (13). While the per capita incidence of SARS-CoV-2 related illness and death has been lower in Canada than in the United States (15, 16), the pandemic has been the cause of tens of thousands of excess deaths in Canada, notwithstanding likely under-counting of SARS-CoV-2 attributable mortality (17). A multi-province Canadian surveillance group has reported elevated risk of hospitalization and critical illness associated with SARS-CoV-2 in pregnant individuals in Canada, but this report was published in early June 2021, just as the Delta variant of concern (VOC) was emerging, and relatively early in the Canadian SARS-CoV-2 vaccination effort (18, 19). Furthermore, this report was largely descriptive, and did not adjust for confounding by VOC, vaccination, age, underlying comorbidity, or healthcare worker status.

The Canadian province of Ontario represents a large (population 14.6 million) and diverse jurisdiction, with high levels of SARS-CoV-2 vaccine coverage (approximately 80% as of December 2021) (20, 21). Ontario has weathered multiple pandemic waves due to the original circulating variant of SARS-CoV-2, and latterly waves caused by the Alpha (spring 2021) and Delta (summer and autumn, 2021) VOC (22), with the Omicron VOC displacing Delta in December 2021 (23).

The province’s rich data sources provide an opportunity for evaluation of the impact of SARS-CoV-2 in pregnant women, relative to other women of childbearing age, and with adjustment for important confounders. Our principal objective was to evaluate the relative morbidity associated with SARS-CoV-2 infection in pregnancy in Ontario. Secondary objectives were to evaluate the impact of infecting VOC and vaccination status on SARS-CoV-2 outcomes.

## Methods

### Data Sources

As the likelihood of vaccination, the dominant SARS-CoV-2 variant, and the provincial public health response changed over time, we created a time-matched cohort of women infected with SARS-CoV-2 in pregnancy; non-pregnant controls were individuals with SARS-CoV-2 matched to pregnant women by date of positive laboratory test for SARS-CoV-2. Pregnant individuals and matched non-pregnant controls were identified in the Province’s Case and Contact Management (CCM) database as described elsewhere (24, 25); we included only cases with a unique “pseudo-health card number”, which permitted linkage with the provincial vaccination database. Control selection was limited to women of reproductive age. As data were available by 10-year age bands, we considered “reproductive age” to encompass individuals aged 10 to 49. Data on comorbidities, healthcare worker status, and infecting variant were also available in CCM. Comorbidities included neurologic disorders, asthma, renal conditions, blood disorders, cancer, cardiovascular conditions, liver conditions, and obesity. We included the time period between January 1, 2020 and January 4, 2022 in the analysis.

Population denominators for the population overall and for women aged 10-49 in Ontario, were obtained from Statistics Canada (20). We estimated person-time at risk of SARS-CoV-2 infection among pregnant individuals based on reports of 107,855 live births and 482 stillbirths among women of childbearing age in Ontario between January 1 and October 31, 2021 (26). Person time at risk was adjusted based on an assumed duration of pregnancy of 40 weeks. To estimate person-time at risk of SARS-CoV-2 infection for non-pregnant women of childbearing age, we subtracted estimated annual person time at risk among pregnant individuals from the population of women of childbearing age. The person-time at risk among those not classified as women of childbearing age was estimated as the total population size, minus the population of women of childbearing age. Variants were classified as non-variant of concern, N501Y+ variant (including the Alpha, Beta and Gamma variants), or Delta variant, as described elsewhere (25). Individuals were considered infected with the Omicron variant (B.1.1.529) if they had been identified as such through viral sequencing, if they were infected with a strain with S-gene target failure on PCR, or with the N501Y mutation, on or after November 10, 2021.

Vaccination information on cases and controls was extracted from the Province’s COVaxON dataset (25), which includes dosage dates and vaccines used. To account for time to develop immunity, we considered individuals to have been vaccinated with a first dose of vaccine during time at risk 14 or more days after the date of their first vaccine dose; individuals were considered vaccinated with two doses of vaccine during time at risk 14 or more days after their second vaccine dose. A flow diagram outlining creation of the cohort is presented in **Figure 1**.

**Figure 1.**
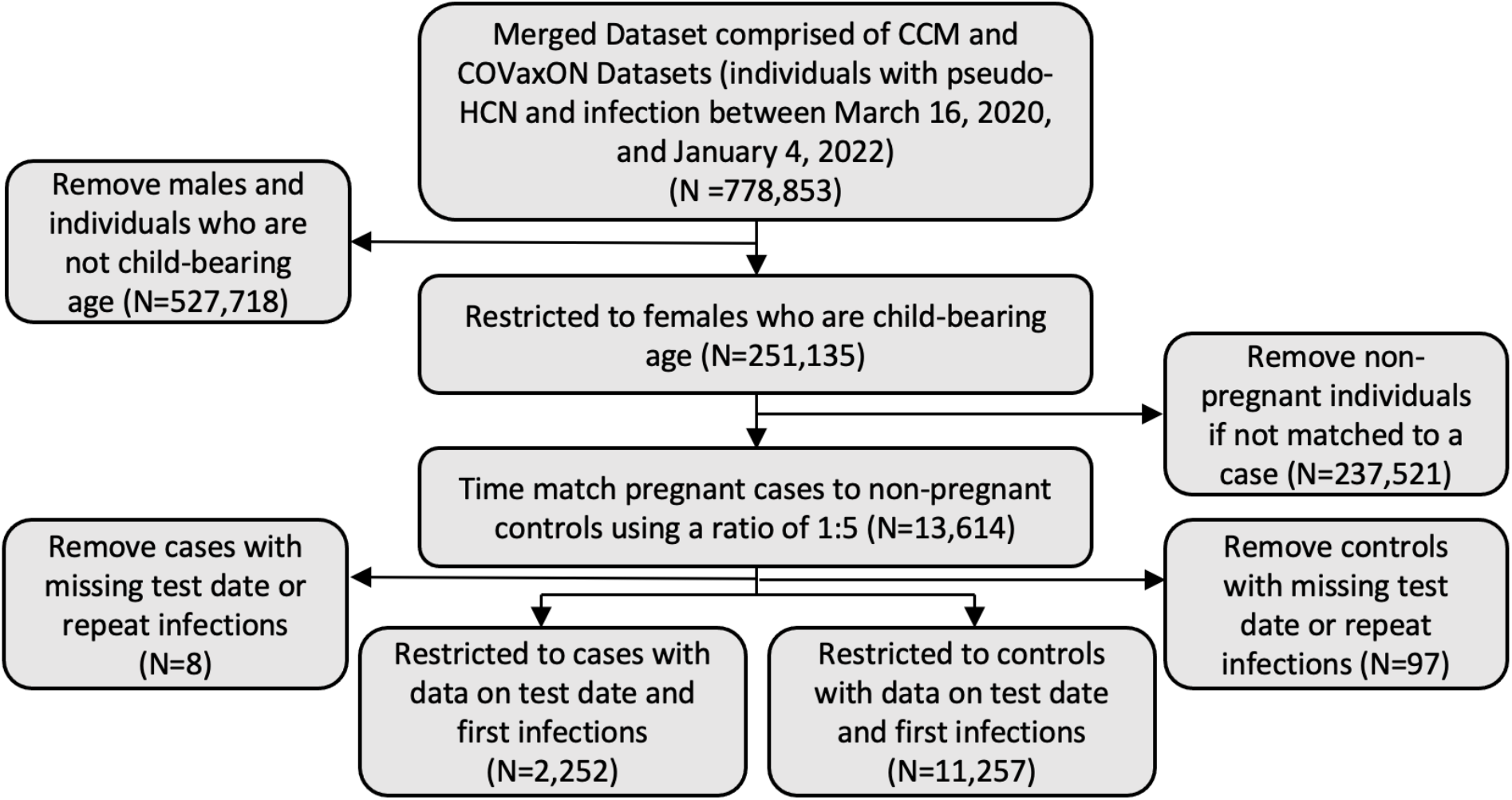
Flow diagram for creation of case-cohort sample. **NOTE:** CCM, Ontario’s COVID-19 Contact Case Management Database; COVaxON, Ontario’s COVID-19 vaccination database.

### Analysis

We explored temporal trends in case incidence overall, in women aged 10-49, and in pregnant women, graphically and through calculation of standardized morbidity ratios (SMR) as described in (27). Briefly, SMR were estimated as incidence in pregnant women, or non-pregnant women of child-bearing age, divided by incidence in the population overall. We calculated SMR for the entire study period, and by week. Confidence intervals for proportions were estimated based on standard errors for log-SMR as in (27).

We used multivariable conditional logistic regression models to estimate the risk of severe illness among our matched cohort while adjusting for age (treated as a four-level ordinal variable), comorbidity, healthcare worker status, vaccination status, and infecting variant, all of which were selected a priori. Severe illness was defined as hospitalization or intensive care unit (ICU) admission. We were not able to include death as an outcome because fewer than five pregnant individuals had died of SARS-CoV-2 in Ontario at the time of analysis. Due to small numbers of adverse outcomes in individuals vaccinated with three doses, we treated vaccination as an ordinal variable with three levels (i.e., unvaccinated, one dose, and two or more doses).

As some pregnant individuals infected with SARS-CoV-2 might be hospitalized for monitoring, we performed an additional restriction analysis, in which risk of ICU was evaluated among pregnant and non-pregnant women admitted to hospital. We hypothesized that if pregnant individuals were admitted for precautionary reasons, they would be less likely to be admitted to ICU than non-pregnant controls, and if admission was due to severity of illness, pregnant individuals should be as or more likely than non-pregnant controls to be admitted to ICU. Due to lack of within-stratum variance in healthcare worker status and infecting variant among those admitted to hospital, we adjusted only for age, comorbidity, and vaccination in analyses restricted to hospitalized individuals.

We performed a series of exploratory restriction analyses within each stratum of comorbidity status, age group, vaccination status, or infecting variant variables to evaluate modification of the observed effects by these covariates in pregnant individuals and non-pregnant controls. As conditional logistic regression models failed to converge for some of these models, we used logistic regression models with time modeled as a cubic term. Heterogeneity in adjusted odds ratios for hospitalization and ICU admission was evaluated using meta-analytic techniques (i.e., graphically using forest plots, and statistically using the meta-analytic Q statistic). Our study was conducted in accordance with the STROBE guidelines for observational research (28), and received ethics approval from the Research Ethics Board at the University of Toronto.

## Results

Our final cohort consisted of 2,252 pregnant women and 11,257 non-pregnant controls, with 5 controls for all but 3 cases (which had 4 matched controls each), with test dates between March 16, 2020, and January 4, 2022. While the temporal pattern of infection risk in pregnant individuals and non-pregnant women of child-bearing age mirrored risk in the population as a whole over time, risk was elevated in non-pregnant women of childbearing age (SMR 1.28) and decreased in pregnant women (SMR 0.43) (**Figure 2**).

**Figure 2.**
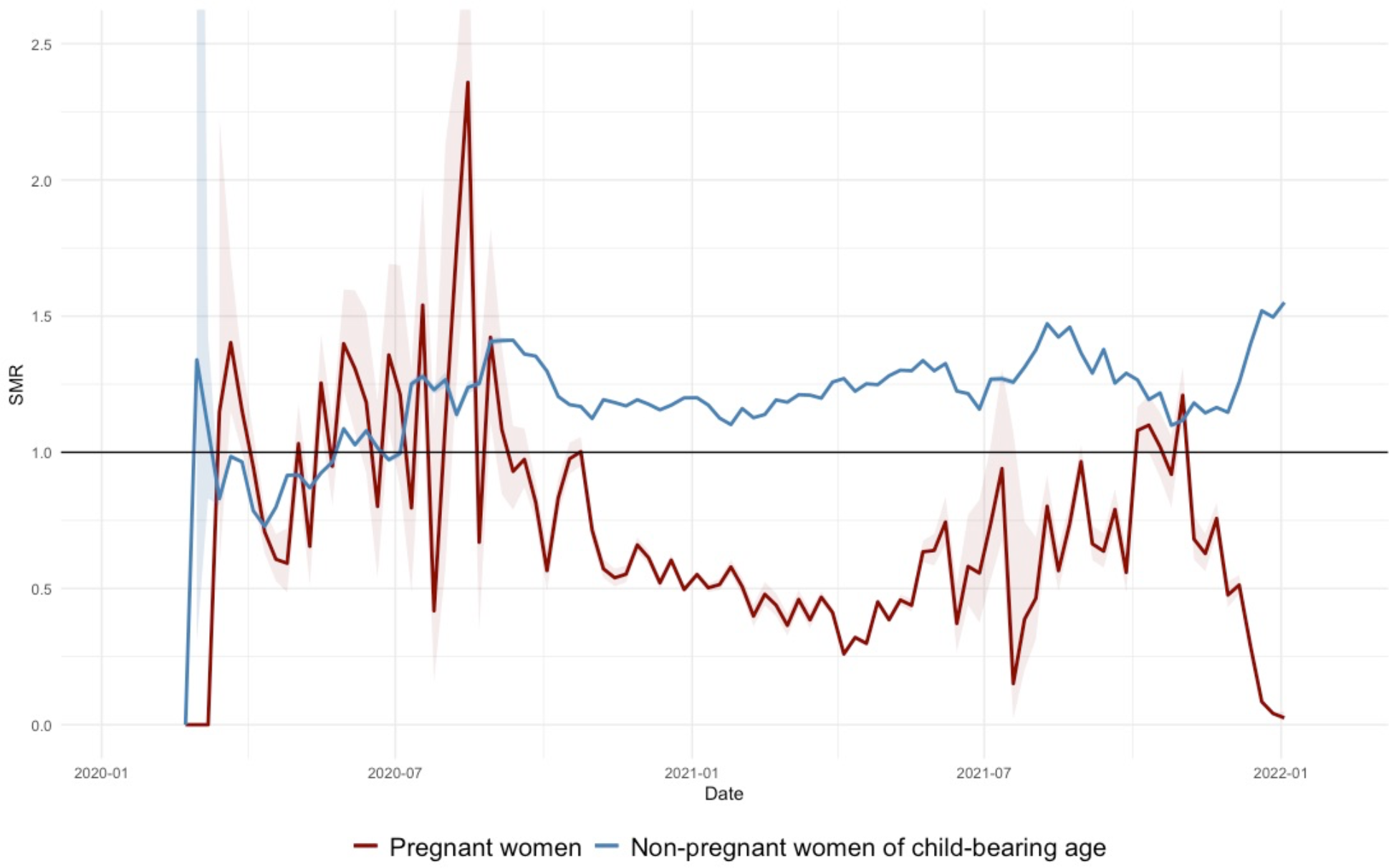
Standardized mortality ratios over time of SARS-CoV-2 cases in pregnant women and non-pregnant women of child-bearing age, Ontario, Canada, January 2020 – January 2022. **NOTE:** Line of reference at (black) depicts overall Ontario population; SMR, standardized mortality ratio.

In our matched cohort, pregnant individuals and non-pregnant controls differed significantly according to risk of hospitalization and ICU admission, as well as age distribution, vaccination status, healthcare worker status, and the presence of any significant comorbidity. Pregnant individuals were more likely than non-pregnant controls to have asthma, diabetes, or a diagnosed hematological disorder. There were no differences between pregnant women and non-pregnant controls with respect to infecting variant, likely because we created a time-matched cohort (**Table 1**).

**Table 1.**
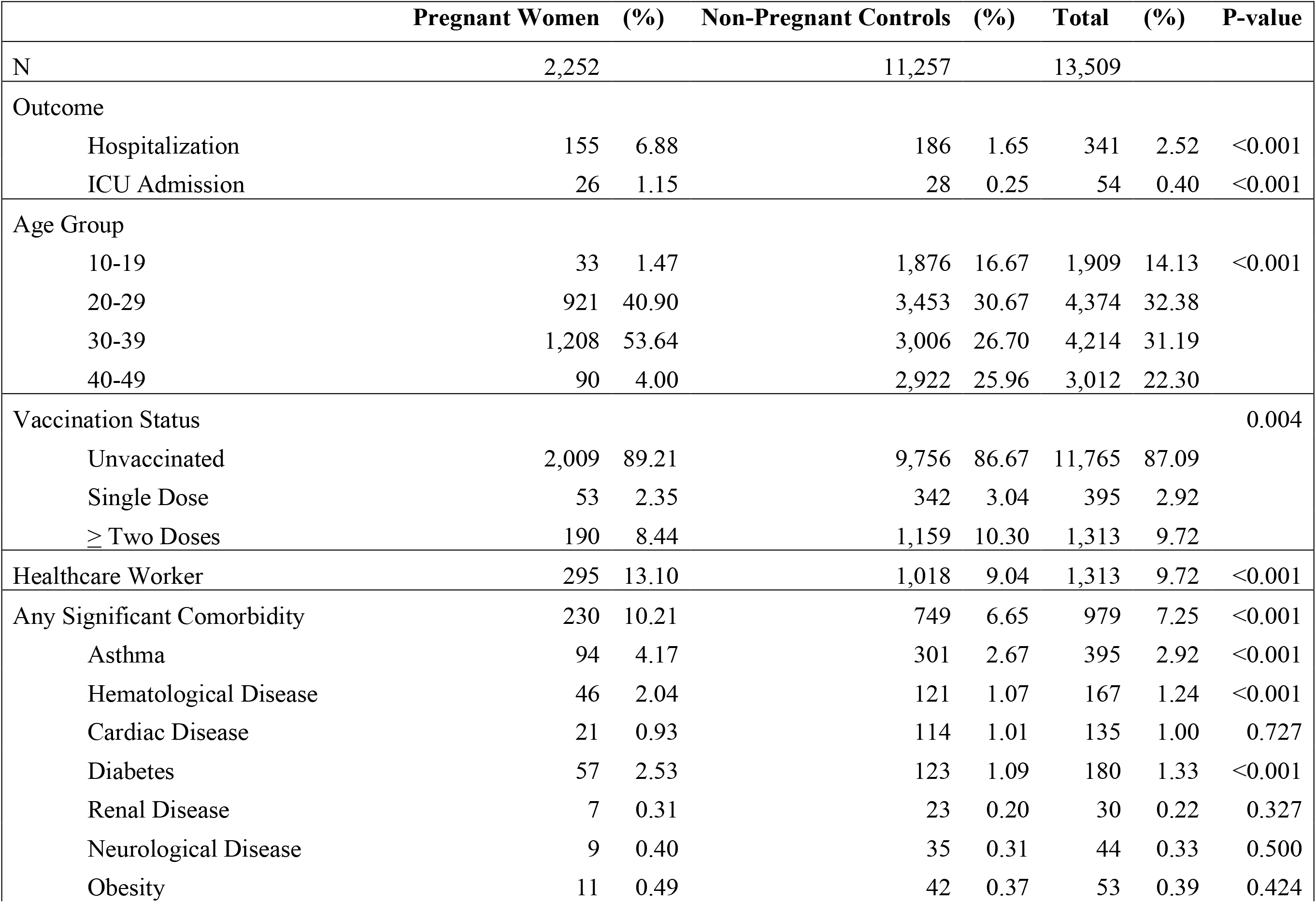
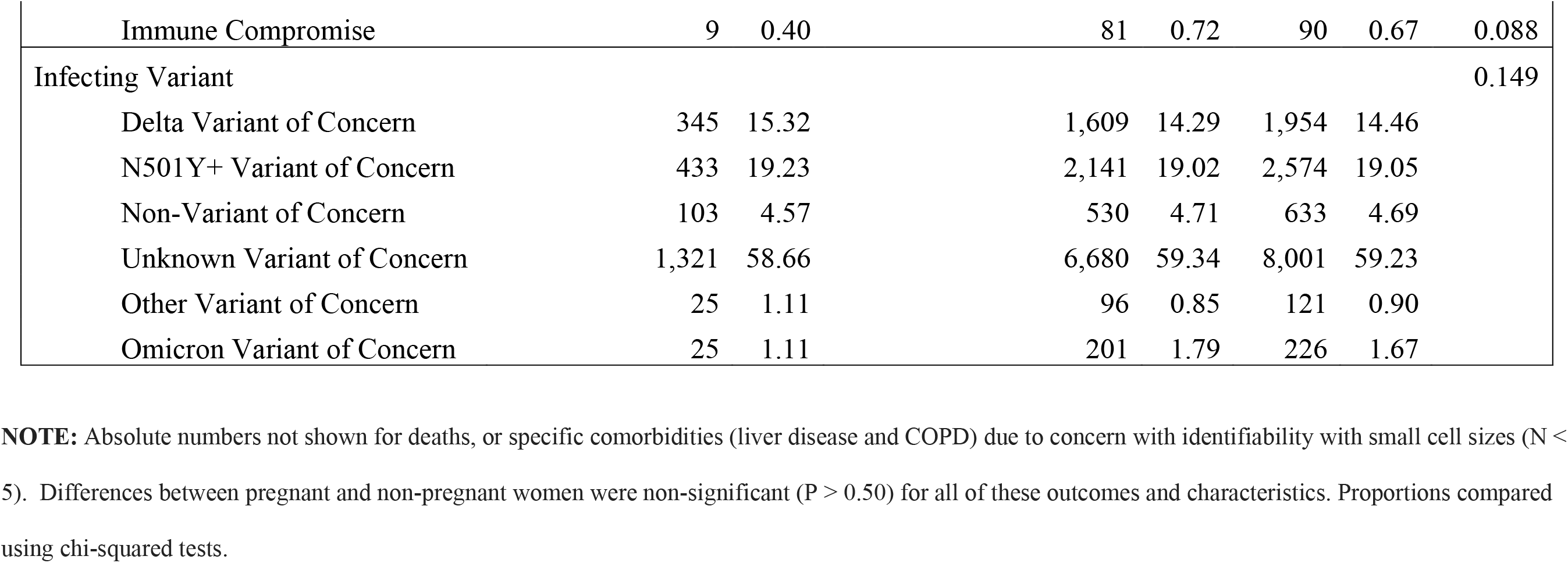
Characteristics of matched study cohort by pregnancy status, Ontario, Canada.

Conditional logistic regression models for hospitalization and ICU admission are presented in **Table 2**. After adjusting for potential confounders, we identified a marked increase in risk of admission to hospital (adjusted OR (aOR) 4.94, 95% CI 3.85 to 6.34) and ICU admission (aOR 6.58, 95% CI 3.29 to 13.18) in pregnant individuals with SARS-CoV-2 infection as compared to non-pregnant controls. We found no significant difference in risk of ICU admission between pregnant and non-pregnant women conditional on hospital admission after adjusting for comorbidity, age, vaccination status, and infecting variant (aOR = 1.30, 95% CI 0.70 to 2.45).

**Table 2.** Results of multivariable conditional logistic regression model on hospitalization and ICU admission in pregnant women due to SARS-CoV-2 infection.

| **Covariate** | **Adjusted Odds Ratio** | **LCL** | **UCL** | **P-value** |
| --- | --- | --- | --- | --- |
| **Hospitalization** |  |  |  |  |
| Pregnancy | 4.94 | 3.85 | 6.34 | <0.001 |
| Vaccination |  |  |  |  |
| Unvaccinated (Referent) | 1.00 |  |  |  |
| 1 Dose | 0.51 | 0.21 | 1.24 | 0.137 |
| 2+ Doses | 0.07 | 0.02 | 0.23 | <0.001 |
| Age Group (per 10-year increase) | 1.69 | 1.45 | 1.98 | <0.001 |
| Healthcare Worker | 0.34 | 0.20 | 0.58 | <0.001 |
| Comorbidity | 3.59 | 2.39 | 5.38 | <0.001 |
| Infecting Variant* |  |  |  |  |
| Delta VOC | 0.86 | 0.47 | 1.59 | 0.638 |
| N501Y+ VOC | 0.82 | 0.50 | 1.34 | 0.430 |
| **ICU Admission** |  |  |  |  |
| Pregnancy | 6.58 | 3.29 | 13.18 | <0.001 |
| Vaccination |  |  |  |  |
| Unvaccinated (Referent) | 1.00 |  |  |  |
| One Dose | 0.34 | 0.02 | 4.87 | 0.424 |
| ≥ Two Doses | 0.25 | 0.02 | 2.55 | 0.241 |
| Age Group | 1.71 | 1.11 | 2.64 | 0.015 |
| Healthcare Worker | 0.09 | 0.02 | 0.47 | 0.005 |
| Comorbidity | 8.71 | 2.88 | 26.36 | <0.001 |
| Infecting Variant* |  |  |  |  |
| Delta VOC | 13.50 | 0.53 | 343.19 | 0.115 |
| N501Y+ VOC | 0.94 | 0.25 | 3.45 | 0.920 |

**NOTE:** ICU, intensive care unit; LCL, 95% lower confidence limit; UCL, 95% upper confidence limit; VOC, variant of concern.
\*Infection with strains classified as non-VOC, other VOC status (including Omicron VOC), or VOC status unknown used as referent.

We performed unmatched logistic regression analyses, with adjustment for time trends and models restricted to individuals with similar comorbidity, vaccination or healthcare worker status, or age group. A summary of the heterogeneity in effect sizes for hospitalizations and ICU admission based on comorbidity status is displayed in **Figure 3**. The odds of hospitalization among pregnant individuals were significantly greater in analyses restricted to those without recorded comorbidities (aOR 5.59, 95% CI 4.34 to 7.20), compared to analyses restricted to those with comorbidities (aOR 2.26, 95% CI 1.28 to 3.99) (P for heterogeneity 0.029), but no heterogeneity in ICU admission risk by presence of comorbidity **(Supplementary Table 1)**. We did not identify heterogeneity in the effect of pregnancy by infecting variant or vaccination status.

**Figure 3.**
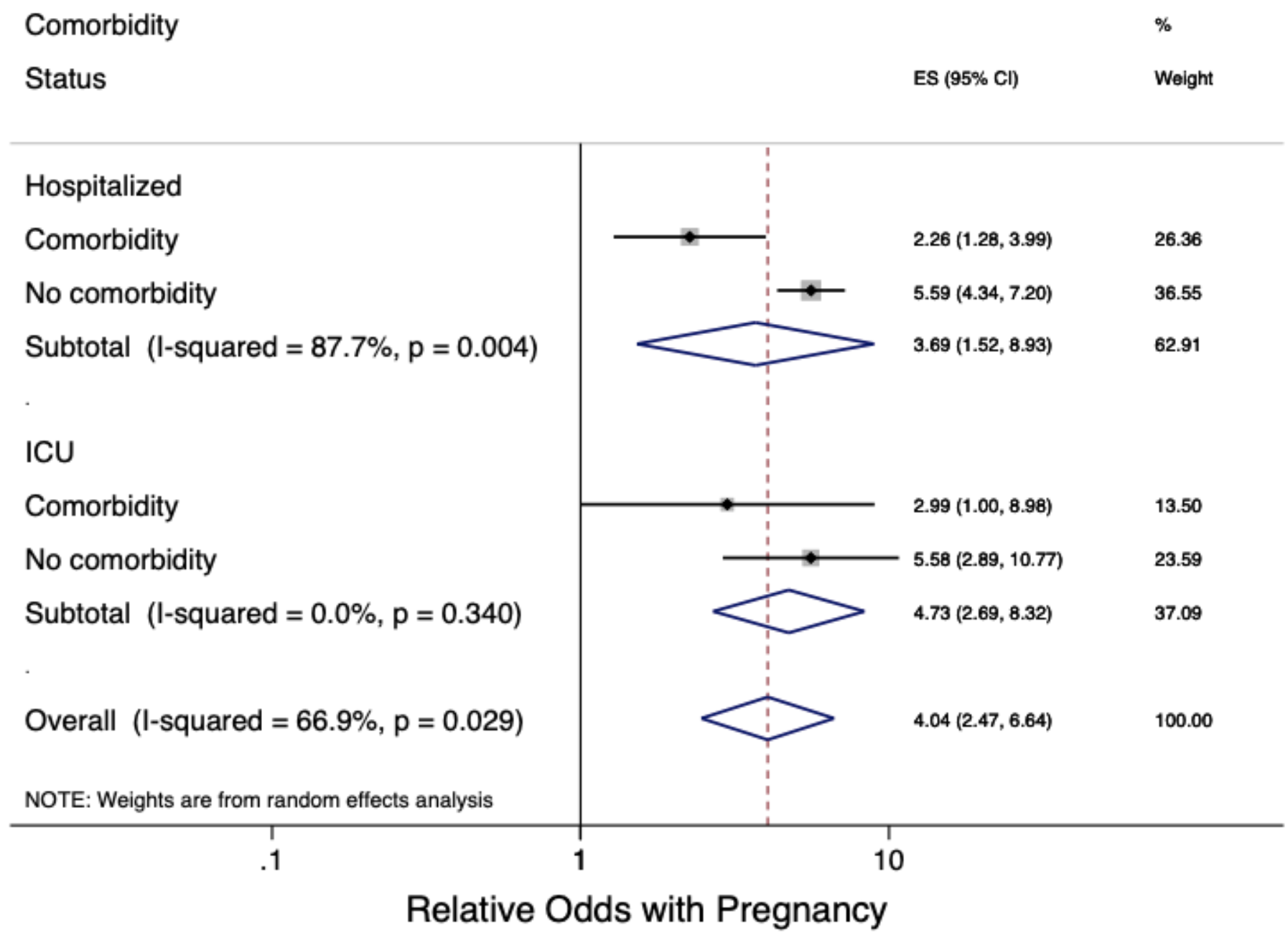
Forest plot summarizing heterogeneity in effect sizes between comorbidity status and severe outcome. **NOTE:** Results are stratified on outcome (hospitalization and ICU admission).

## Discussion

In a cohort of pregnant and non-pregnant women in Ontario, Canada, pregnancy was associated with a decreased risk of infection, but a markedly increased risk of severe illness following infection with SARS-CoV-2. This large increase in risk persisted after adjustment for age, comorbidity, vaccination status, and infecting variant. The fact that pregnant and non-pregnant women were matched by approximate date of infection makes it unlikely that the large increase in hospitalization and ICU risk associated with pregnancy is due to varying propensity to receive vaccination over time, or by temporal changes in circulating variants of concern. Furthermore, we did not see a diminished risk of intensive care admission in hospitalized pregnant women when they were compared to other hospitalized women, suggesting that pregnant women were likely to have been admitted to hospital for severity of respiratory illness rather than simply for monitoring.

The excess risk associated with pregnancy was less pronounced when analyses were restricted to pregnant and non-pregnant women who had comorbidities, again suggesting that in otherwise healthy women, pregnancy itself is a factor that increases illness severity, while in women with comorbidities it becomes one of several factors that augments risk. The physiology of increased severity of respiratory virus infection in pregnancy is complex, and not fully understood in the context of SARS-CoV-2. While pregnant women have increased cardiac demand, diminished pulmonary reserve, and physiological impairment of the immune response (which serves to prevent rejection of the developing fetus), and these changes have been linked to worse outcome with pulmonary infection (5, 29), COVID-19 is not simply an infection of the respiratory system. Numerous vascular and hematological abnormalities occur following SARS-CoV-2 infection, and reports of elevated risk of stillbirth in infected individuals (12), and pathological changes reported in the placenta (30), may suggest that some of the excess morbidity we describe here is related to vascular and hematological disease rather than respiratory disease.

While our primary aim in this study was to evaluate the impact of pregnancy on risk of severe illness with SARS-CoV-2, vaccination, including partial vaccination, was associated with a marked reduction in hospital admission risk and ICU admission in multivariable analyses, notwithstanding the fact that in this case-only analysis, all vaccinated women had, by definition, experienced breakthrough infections. Again, given the safety of these vaccines in pregnancy (8), and the markedly elevated risk of severe illness in pregnancy, risk calculus strongly favours vaccination for pregnant women.

Like any observational study, ours is subject to several limitations. First, the inability to perform linkage of mothers and infants using available data prevents us from evaluating the question of whether maternal vaccination is likely to reduce risk associated with SARS-CoV-2 infection in live-born infants, though anti-spike antibody is present in infants born to vaccinated mothers, and is present at higher titres, and persists longer, than antibody in infants born to mothers with SARS-CoV-2 infection (31). The recent emergence of the Omicron variant makes us unable to explore the relative virulence of this variant in pregnancy in the current paper (23). Our estimates may also be subject to residual confounding by incompletely ascertained factors, including presence of underlying medical conditions. However, the very large effect size associated with pregnancy means that the magnitude of effect of putative confounding factors needed to explain away these associations would be implausibly large (32).

In summary, we identify a large increase in risk of hospitalization and ICU admission in pregnant women infected with SARS-CoV-2 virus, relative to female controls of child-bearing age. This effect was not explained by comorbidity or vaccination status, and indeed, the relative increase in risk with pregnancy was greater when we restricted our analyses to women without medical comorbidities. Vaccination markedly reduced hospitalization and ICU admission risk in all women, pregnant and non-pregnant, in this study, and should be strongly encouraged in pregnancy.

## Data Availability

Our study was conducted in accordance with the STROBE guidelines for observational research, and received ethics approval from the Research Ethics Board at the University of Toronto.

## Acknowledgements

The authors with to thank the staff at Public Health Ontario and Ontario’s public health units for collecting, sequencing, analyzing, and providing access to the data used for this analysis.

## Supplementary Table

**Table S1:** Adjusted odds ratios and 95% confidence intervals from logistic regression on hospitalization and intensive care unit admission due to SARS-COV-2 in pregnant females, stratified by comorbidity status.

| **Outcome and Comorbidity Status** | **aOR\*** | **Lower CI** | **Upper CI** | **P-value** |
| --- | --- | --- | --- | --- |
| Hospitalization |  |  |  |  |
| Comorbidity | 2.26 | 1.28 | 3.99 | 0.005 |
| No Comorbidity | 5.59 | 1.52 | 8.93 | <0.001 |
| ICU Admission |  |  |  |  |
| Comorbidity | 2.99 | 1.00 | 8.98 | 0.050 |
| No Comorbidity | 5.58 | 2.89 | 10.77 | <0.001 |

\*Adjusted for vaccination status, age, healthcare worker status, variants of concern, and time cubed.
**NOTE:** CI, confidence interval; aOR, adjusted odds ratio.

## References

1. Jamieson DJ, Theiler RN, Rasmussen SA. Emerging infections and pregnancy. Emerg Infect Dis. 2006;12(11):1638–43.

2. Szekeres-Bartho J. Immunological relationship between the mother and the fetus. Int Rev Immunol. 2002;21(6):471–95.

3. Maestre A, Carmona-Fonseca J. Immune responses during gestational malaria: a review of the current knowledge and future trend of research. J Infect Dev Ctries. 2014;8(4):391–402.

4. Krain LJ, Nelson KE, Labrique AB. Host immune status and response to hepatitis E virus infection. Clin Microbiol Rev. 2014;27(1):139–65.

5. Bhatia PK, Biyani G, Mohammed S, Sethi P, Bihani P. Acute respiratory failure and mechanical ventilation in pregnant patient: A narrative review of literature. J Anaesthesiol Clin Pharmacol. 2016;32(4):431–9.

6. Pacheco LD, Ghulmiyyah L. Ventilator management in critical illness. Available via the Internet at https://books.scholarsportal.info/en/read?id=/ebooks/ebooks2/wiley/2011-12-13/4/9781444316780. Last accessed December 27, 2021. In: Belfort MA, Saade G, Foley MR, Phelan JP, Dildy GA, editors. Critical Care Obstetrics. 5 ed. Oxford, UK: Wiley-Blackwell; 2010. p. 124–51.

7. Yankowitz JS M., Fetal effects of drugs commonly used in critical care. Available via the Internet at https://books.scholarsportal.info/en/read?id=/ebooks/ebooks2/wiley/2011-12-13/4/9781444316780. Last accessed December 27, 2021. In: Belfort MA, Saade G, Foley MR, Phelan JP, Dildy GA, editors. Critical Care Obstetrics. 5 ed. Oxford, UK: Wiley-Blackwell; 2010. p. 626–38.

8. Fu W, Sivajohan B, McClymont E, Albert A, Elwood C, Ogilvie G, et al. Systematic review of the safety, immunogenicity, and effectiveness of COVID-19 vaccines in pregnant and lactating individuals and their infants. Int J Gynaecol Obstet. 2021.

9. Geoghegan S, Stephens LC, Feemster KA, Drew RJ, Eogan M, Butler KM. “This choice does not just affect me.” Attitudes of pregnant women toward COVID-19 vaccines: a mixed-methods study. Hum Vaccin Immunother. 2021;17(10):3371–6.

10. Riad A, Jouzova A, Ustun B, Lagova E, Hruban L, Janku P, et al. COVID-19 Vaccine Acceptance of Pregnant and Lactating Women (PLW) in Czechia: An Analytical Cross-Sectional Study. Int J Environ Res Public Health. 2021;18(24).

11. Johns Hopkins University Coronavirus Resource Centre. COVID-19 Dashboard by the Center for Systems Science and Engineering (CSSE) at Johns Hopkins University (JHU). Available via the Internet at https://coronavirus.jhu.edu/map.html. Last accessed December 27, 2021.. 2021.

12. DeSisto CL, Wallace B, Simeone RM, Polen K, Ko JY, Meaney-Delman D, et al. Risk for Stillbirth Among Women With and Without COVID-19 at Delivery Hospitalization -United States, March 2020-September 2021. MMWR Morb Mortal Wkly Rep. 2021;70(47):1640–5.

13. Zambrano LD, Ellington S, Strid P, Galang RR, Oduyebo T, Tong VT, et al. Update: Characteristics of Symptomatic Women of Reproductive Age with Laboratory-Confirmed SARS-CoV-2 Infection by Pregnancy Status - United States, January 22-October 3, 2020. MMWR Morb Mortal Wkly Rep. 2020;69(44):1641–7.

14. Ellington S, Strid P, Tong VT, Woodworth K, Galang RR, Zambrano LD, et al. Characteristics of Women of Reproductive Age with Laboratory-Confirmed SARS-CoV-2 Infection by Pregnancy Status - United States, January 22-June 7, 2020. MMWR Morb Mortal Wkly Rep. 2020;69(25):769–75.

15. Ritchie H, Mathieu E, Rodés-Guirao L, Appel C, Giattino C, Ortiz-Ospina E, et al. Canada: Coronavirus Pandemic Country Profile. Available via the Internet at https://ourworldindata.org/coronavirus/country/canada. Last accessed December 27, 2021. 2021.

16. Johns Hopkins University Coronavirus Resource Centre. COVID-19 United States Cases by County. Available via the Internet at https://coronavirus.jhu.edu/us-map. Last accessed December 27, 2021.. 2021.

17. Moriarty T, Boczula AE, Thind EK, Loreto N, McElhaney JE. Excess All-Cause Mortality During the COVID-19 Epidemic in Canada. Available via the Internet at https://rsc-src.ca/sites/default/files/EM%20PB_EN.pdf. Last accessed December 27, 2021. Ottawa, Canada: Royal Society of Canada; 2021.

18. McClymont E, Abenhaim H, Albert A, Boucoiran I, Cassell K, Castillo E, et al. Canadian Surveillance of COVID-19 in Pregnancy (CANCOVID-Preg): A Rapidly Coordinated National Response Using Established Regional Infrastructures. J Obstet Gynaecol Can. 2021;43(2):165–6.

19. Money D. Canadian surveillance of COVID-19 in pregnancy: Epidemiology, maternal and infant outcomes. Available via the Internet at http://med-fom-ridprogram.sites.olt.ubc.ca/files/2021/06/CANCOVID_Preg-Report-4-ON-BC-QC-MB-AB_FINAL.pdf. Last accessed December 27, 2021.; 2021.

20. Statistics Canada. Population estimates, quarterly. Table: 17-10-0009-01 (formerly CANSIM 051-0005). Available via the Internet at https://www150.statcan.gc.ca/t1/tbl1/en/tv.action?pid=1710000901. Last accessed May 29, 2020. 2020.

21. Ontario Agency for Health Protection and Promotion (Public Health Ontario). COVID-19 vaccine uptake in Ontario: December 14, 2020 to November 28, 2021. Available via the Internet at publichealthontario.ca/-/media/documents/ncov/epi/covid-19-vaccine-uptake-ontario-epi-summary.pdf?la=en. Last accessed December 7, 2021. 2021.

22. Fisman DN, Tuite AR. Progressive Increase in Virulence of Novel SARS-CoV-2 Variants in Ontario, Canada. medRxiv. 2021:2021.07.05.21260050.

23. Garcia-Beltran WF, St. Denis KJ, Hoelzemer A, Lam EC, Nitido AD, Sheehan ML, et al. mRNA-based COVID-19 vaccine boosters induce neutralizing immunity against SARS-CoV-2 Omicron variant. medRxiv. 2021:2021.12.14.21267755.

24. Fisman DN, Greer AL, Hillmer M, O’Brien SF, Drews SJ, Tuite AR. COVID-19 case age distribution: correction for differential testing by age. medRxiv. 2020:2020.09.15.20193862.

25. Fisman DN, Tuite AR. Evaluation of the relative virulence of novel SARS-CoV-2 variants: a retrospective cohort study in Ontario, Canada. CMAJ. 2021;193(42):E1619–E25.

26. Better Outcomes Registry and Network (BORN) Ontario. Number of Live Births and Stillbirths among Infants born in Ontario, by COVID-19 Vaccination Status. Available via the Internet at https://www.bornontario.ca/en/news/number-of-live-and-stillbirths-among-infants-born-in-ontario-by-covid-19-vaccination-status.aspx. Last accessed January 17, 2022. 2021.

27. Fisman DN, Greer AL, Brankston G, Hillmer M, O’Brien SF, Drews SJ, et al. COVID-19 Case Age Distribution: Correction for Differential Testing by Age. Ann Intern Med. 2021.

28. von Elm E, Altman DG, Egger M, Pocock SJ, Gotzsche PC, Vandenbroucke JP, et al. The Strengthening the Reporting of Observational Studies in Epidemiology (STROBE) statement: guidelines for reporting observational studies. Bull World Health Organ. 2007;85(11):867–72.

29. Tamma PD, Steinhoff MC, Omer SB. Influenza infection and vaccination in pregnant women. Expert Rev Respir Med. 2010;4(3):321–8.

30. Menter T, Mertz KD, Jiang S, Chen H, Monod C, Tzankov A, et al. Placental Pathology Findings during and after SARS-CoV-2 Infection: Features of Villitis and Malperfusion. Pathobiology. 2021;88(1):69–77.

31. Shook LL, Atyeo CG, Yonker LM, Fasano A, Gray KJ, Alter G, et al. Durability of Anti-Spike Antibodies in Infants After Maternal COVID-19 Vaccination or Natural Infection. JAMA. 2022.

32. VanderWeele TJ, Ding P. Sensitivity Analysis in Observational Research: Introducing the E-Value. Ann Intern Med. 2017;167(4):268–74.

